# The effect of national traditional sports on weight loss and blood lipid health of overweight and obese students: Meta analysis

**DOI:** 10.1101/2025.06.13.25329601

**Authors:** Fengling Zhou, Ping Sun

## Abstract

**Background:** The problem of overweight and obesity in children and adolescents around the world is serious, and traditional intervention methods have problems such as poor compliance. Traditional ethnic sports (such as martial arts and tai chi) combined with physical and mental exercise and cultural elements may provide a new way for obesity management, but there is a lack of systematic evaluation.

A systematic literature search was conducted in twelve electronic databases (PubMed, Cochrane Library, Web of Science, MEDLINE, EBSCO, CINAHL, OVID, Embase, Scopus, SPORTDiscus, CNKI, ProQuest) from inception to March 12, 2025. Randomized controlled trials of ethnic traditional sports interventions in overweight or obese students (age 7.6–23 years; BMI > 23.9 kg/m²) were eligible, yielding twelve RCTs (n = 520). Two reviewers independently screened studies, extracted data, and assessed risk of bias using the Cochrane RoB tool.

Across 12 RCTs (n = 520), ethnic traditional sports interventions did not significantly alter body weight (WMD = –0.115 kg; 95% CI: –0.30 to 0.07; p = 0.233) or waist circumference (WMD = –0.34 cm; 95% CI: –0.82 to 0.13; p = 0.156), but yielded significant reductions in BMI (WMD = –0.33 kg/m²; 95% CI: –0.60 to –0.06; p = 0.016) and body fat percentage (WMD = –0.31%; 95% CI: –0.52 to –0.10; p = 0.004). In lipid profiles, high-density lipoprotein cholesterol increased (WMD = 0.33 mmol/L; 95% CI: 0.05 to 0.61; p = 0.020), whereas total cholesterol (WMD = –0.01 mmol/L; p = 0.952), triglycerides (WMD = 0.33 mmol/L; p = 0.468) and low-density lipoprotein cholesterol (WMD = 0.03 mmol/L; p = 0.828) remained unchanged. Subgroup analyses indicated that Chinese traditional exercises (e.g. Tai Chi, Kung Fu) produced more pronounced reductions in BMI and body fat than other modalities.

**Conclusion:** National traditional sports can effectively improve the BMI, body fat rate and HDL-C level of overweight and obese students. It is a potential intervention method. In the future, it is necessary to further standardize the program and expand the research sample.

## 1 Introduction

In recent years, the prevalence of overweight and obesity among children and adolescents has risen markedly worldwide, representing a major public health challenge. According to the World Health Organization, over 340 million individuals aged 5–19 years are currently classified as overweight or obese, with this figure continuing to increase. Obesity not only increases the risk of chronic diseases, but also may affect mental health, causing problems such as inferiority and anxiety [2]. Empirical evidence demonstrates the prevalence of overweight and obesity has a strong association with lifestyle changes, dietary imbalance, lack of exercise and other factors, especially the sedentary and excessive use of electronic equipment in the student population. At present, the main intervention methods such as diet control, aerobic exercise, and resistance training have problems such as poor compliance and lack of interest. As an intervention method that integrates culture, physical and mental exercise and low-impact exercise, national traditional sports has unique advantages. It has unique advantages. Through the combination of ‘meaning, qi and shape’, it can not only improve physical function, but also regulate psychological state and improve exercise compliance [3]. However, most of the existing studies focus on the western sports model, and the research on its effect is controversial. Currently, no systematic meta-analysis has been conducted to assess the effects of traditional ethnic sports on body composition and blood lipid profiles in overweight and obese students. This study aims to investigate the effects of traditional ethnic sports on body composition and blood lipid parameters in overweight studentsthrough Meta-analysis, to fill the existing research gaps, to provide a basis for formulating more effective exercise intervention programs, and to provide reference for public health policy formulation.

## 2 Materials and methods

### 2.1 Search strategy

A comprehensive literature search was conducted across the following databases: PubMed, Cochrane Library, Wob of Science, MEDLINE, EBSCO, CINAHL, OVID, Embase, Scopus, SPORTDiscus, CNKI, ProQuest, FMRS, Cochrane Central Register of Controlled Trials database. The literature search was updated through March 12, 2025, employing the following primary search terms:’ Martial Arts’ (e.g.,’ Judo’,’ Karate’,’ Kung Fu’,’ Tae Kwon Do’,’ Wushu’,’ Hap Ki Do’,’ Tai Chi’, etc.) and’ student’ (e.g.,’ Adolescent’,’ Youths’,’ Teens’,’ Teenagers’,’ Child’,’ Undergraduate’, etc.) and’ Overweight or Obesity’ (for example,’ Body Weight’,’ Abdominal Fat’,’ Central Obesity’,’ Hypernutrition’, etc.). The reference lists of pertinent articles were also reviewed. The research programme is registered with PROSPERO (ID: 420251011934).

### 2.2 Inclusion and exclusion criteria

The study selection process adhered to the Preferred Reporting Items for Systematic Reviews and Meta-Analyses (PRISMA) guidelines (Figure 1). Studies were included according to the following criteria:

(1) The subjects were overweight and obese students; age 7.6-23 years old, BMI > 23.9.
(2) Intervention through national traditional sports, using randomized controlled trials;
(3) In the traditional exercises (including martial arts, Yang’s tai chi, boxing, judo, taekwondo, karate, eight five step tai chi, traditional and style tai chi) before and after the intervention measurement index
(4) Randomized controlled trials;

**Fig. 1.**
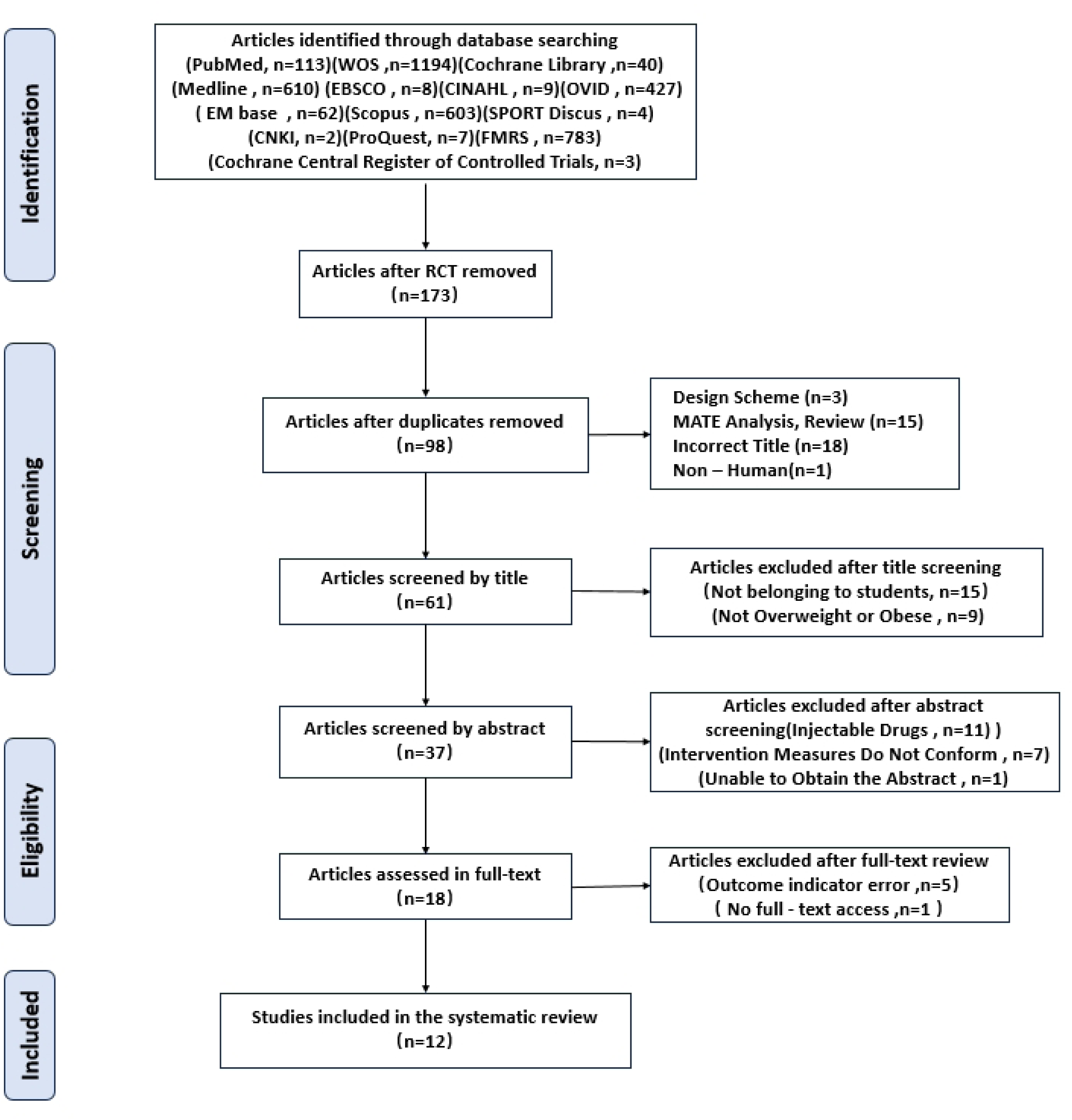
The screening flow chart of the included studies.

Exclusion criteria are as follows:

(1) non-randomized controlled trials, such as semi-randomized controlled trials, animal studies, protocols, meeting abstracts, research protocols, expert letters, editorials, notes and book chapters;
(2) incomplete data or unreported, unable to obtain the full text of the study;
(3) pure descriptive research;
(4) Data or analysis Repeated studies;
(5) Research on non-conformity of intervention objects, intervention methods or outcome indicators (including height, waist circumference, weight, BF, BMI, TC, LDL-C, TG and HDL-C);

### 2.3 Data extraction

A Two independent reviewers screened all eligible articles and performed data extraction. The search cannot perform full-text review only based on articles classified according to title and abstract. Discrepancies were resolved through discussion between the two reviewers and, if necessary, adjudicated by a third reviewer. For each study, the following data were extracted:(1) Basic information of the included literature (title, first author, year of publication, journal);

(2) Subject characteristics (sample size, age, gender, race);
(3) Exercise intervention (exercise type, intensity, duration, frequency, intervention time);
(4) Control group intervention measures;
(5) Relevant information of literature quality evaluation;
(6) Outcome indicators and main findings.

### 2.4 Quality assessment

Two independent reviewers assessed the methodological quality of included studies using the Cochrane Risk of Bias tool as recommended in the Cochrane Handbook (Figures 2 and 3). Six domains—selection bias, performance bias, detection bias, attrition bias, reporting bias, and other biases—were evaluated and rated as low, unclear, or high risk of bias. The overall level of evidence was graded according to GRADE criteria, with all studies initially classified as high-quality evidence. According to bias, inconsistency, indirectness, publication bias or accuracy risk, the level of evidence can be reduced to high, medium, low or very low. The two researchers independently evaluated the level of evidence. If the opinions were inconsistent, the third party discussed and jointly decided the final rating.

**Fig. 2.**
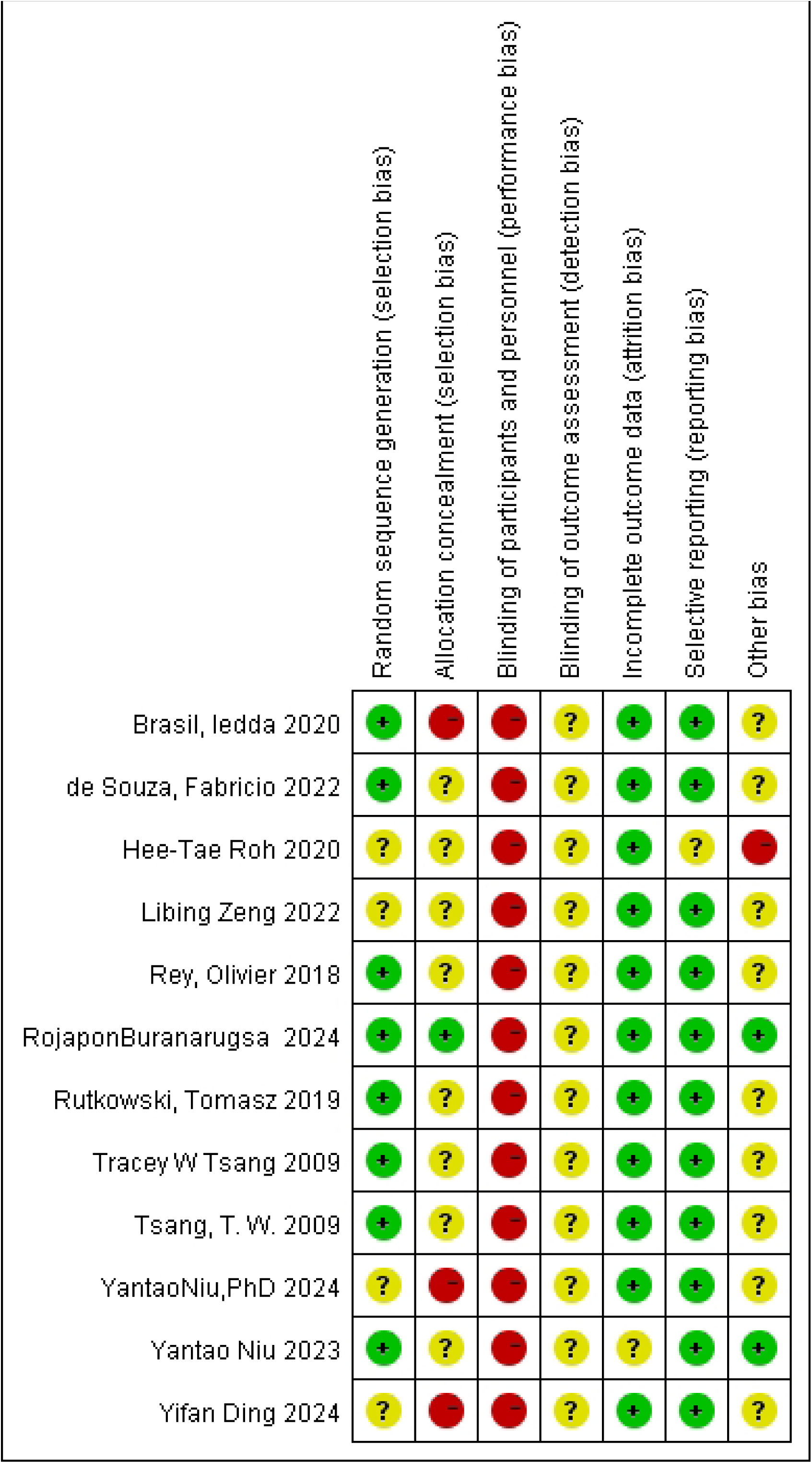
Risk of Bias Summary.

**Fig. 3.**
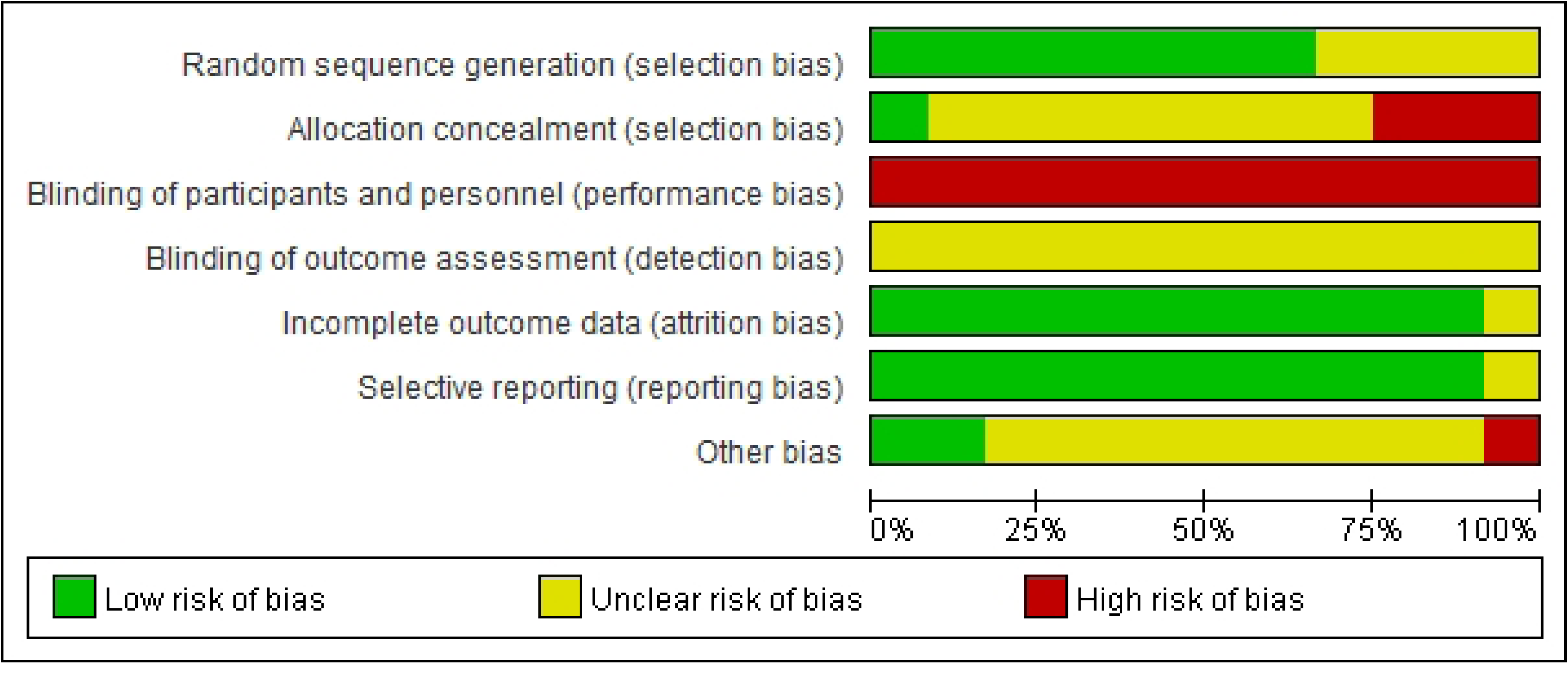
Risk of Bias Graph.

### 2.5 Statistical analysis

This study used Windows Review Manager 5.3.0 version and STATA 18.0 version software for meta-analysis. If the research results are considered to be homogeneous, they are merged. Table 1 lists the measurements used in the included studies, and all outcome indicators are continuous variables. Data were extracted as mean differences with corresponding standard deviations from baseline to post-intervention for each group. Pooled effect sizes were calculated using weighted mean differences (WMDs) or standardized mean differences (SMDs), each presented with 95% confidence intervals (CIs). When the p value of the Q test is 0.05, the comprehensive effect is statistically significant. The random effects model was used for meta-analysis, and the I^2^ test was used to evaluate statistical heterogeneity. The I^2^ value represents the percentage of variation in the estimated effect. When the I^2^ value is 25 %, 50 % and 75 %, it represents low, medium and high heterogeneity, respectively. When I^2^<50 % and P>0.1, the fixed effect model is used to combine the effect size; when I^2^ > 50 % and P< 0.1, a random effect model was used to explore the source of heterogeneity through subgroup analysis and Meta regression analysis. When substantial heterogeneity is detected, sensitivity analyses should be conducted by sequentially excluding individual studies to identify sources of heterogeneity and assess their impact on the overall results. Finally,the estimated value of the processing difference is presented in the forest map.

**Table 1.** Main characteristics of all studies included in the Mate analysis.

| Study cohort | Year | Study region | Ethnicity | NO.(M/F) | Follow-up(moths)<br>(median and range) | Intervention | Age (years;<br>median and range) | Outcome | Type | HR | RoB Tool |
| --- | --- | --- | --- | --- | --- | --- | --- | --- | --- | --- | --- |
| Tsang, T. W. | 2009 | Australia | Caucasian | 20(8/12) | 6 | KF/TC | 13.5 ( 11-16 ) | LDL-C; TG; HDL-C; TC | RCT | R(M/U) | MRoB |
| Yifan Ding | 2024 | China | Asian | 40(NR) | 12 | KF | 16.5 ( 15-18 ) | BMI; LDL-C; TG; HDL-C; BW; WC; Height; ST; TC | RCT | R(U) | MRoB |
| Libing Zeng | 2022 | China | Asian | 50(25/25) | 4 | KF | 14.5 ( 12-17 ) | BMI;BF;BW;WC ; | RCT | R(M/U) | MRoB |
| Rey, Olivier | 2018 | USA | Caucasian | 24(10/14 ) | 5 | Boxing | 14.6 | BF;BW;Height ; | RCT | R(M) | MRoB |
| Tracey W Tsang | 2009 | Australia | Caucasian | 20(8/12) | 6 | KF | 9 ( 6-12 ) | BMI;BF;BW;WC;Height ; | RCT | R(M/U) | MRoB |
| Rojapon Buranarugsa | 2024 | Thailand | Asian | 81(81/0) | 12 | BW-TC; TH-TC | 18-23 | Height; ST; BW | RCT | R(M) | MRoB |
| Brasil, Iedda | 2020 | Brazil | Caucasian | 35 ( 17/18 ) | 12 | Judo | 10.5 ( 8-13 ) | BMI;BF;BW;WC;Height ; | RCT | R(M/U) | MRoB |
| de Souza, Fabricio | 2022 | Brazil | Caucasian | 70(30/40) | 12 | Karate | 14.5 ( 12-17 ) | BMI; LDL-C; TG; HDL-C; BF; BW; Height; TC | RCT | R(M/U) | MRoB |
| Hee-Tae Roh | 2020 | Korea | Asian | 20(NR) | 16 | Taekwondo | 12.55±0.51 | BMI; BW; Height; | RCT | R(M/U) | MRoB |
| Yantao Niu | 2023 | China | Asian | 81(81/0) | 12 | BW-TC; TH-TC | 20.5(18-23) | LDL-C; HDL-C; Height; BW; TC; BF; | RCT | R(M/U) | MRoB |
| Rutkowski, Tomasz | 2019 | Poland | European | 59(30/29) | 10 | karate | 7.6±0.4 | BMI; BF; BW; Height; | RCT | R(U) | MRoB |
| YantaoNiu, PhD | 2024 | China | Asian | 20(20/0) | 2 | BW-TC; TH-TC | 18-23 | BMI; BW; Height; ST; | RCT | R(U) | MRoB |

KF : Kung Fu ; TC:Yang style Tai Chi;BW-TC:Bafa Wubu Tai Chi ; TH-TC: traditional He-style Tai Chi; R."M"means the HRcome fom mulianiale anabsis,"u" means theHR comes from univanale
analysis; BW: Body weight ; ST: Sit-and-reach test; WC: Waist circumference; TC: Totalcholesterol; RoB Tool: Cochrane Risk of Bias Tool ; MRoB: Moderate Risk of Bias

## 3 Results

### 3.1 Screening results and research characteristics

A total of 3865 articles were screened out by the initial search strategy. After screening, 12 studies were finally included, involving 520 students, and the research time span was from 2009 to 2024 (Fig.1). Six of the participants were Asian [ 5,6,9,12,13,15], five were white [ 4,7,8,10,11], and one was European [14]. Research sources include China (4 studies) [ 5,6,13,15], Australia [4,8] and Brazil (2 studies each) [10,11], as well as the United States [7], South Korea [12], Thailand [9] and Poland [14] (1 study each). Twelve studies reported HR directly, of which 10 were calculated by univariate analysis and 9 by multivariate analysis. There were 7 queues with less than 50 participants [ 4,5,7,8,10,12,15] and 5 queues with more than 50 participants [ 6,9,11,13,14]. The results of the study covered 9 indicators, 10 related to height [5,7–9,10–15], 4 related to waist circumference [5,6,8,10], 11 related to weight [5–15], 7 related to BF [ 3–8,10,11,13,14], 4 related to TC [ 4,5,11,12], 8 related to BMI [ 5,6,8,10–12,14,15], 4 related to LDL-C [ 4,5,11,13], 3 related to TG [4,5,11], 4 related to HDL-C [4,5,11,13].All included studies examined the association between traditional ethnic sports and outcomes in overweight and obese students. Detailed study characteristics are presented in Table 1.

### 3.2 Quality evaluation of the included literature

The 12 RCTs were evaluated according to the 6 areas of the bias risk assessment tool ROB2.0. Figure 2 and Figure 3 show the details of the evaluation. When assessing the risk of bias in the random process, four RCTs were rated as potential risks because they did not describe whether the allocation order was hidden. When assessing the intervention bias, 1 RCT was at a low risk of bias [9], 8 RCTs may have a risk of bias [4,7–8,11–14], and the intervention bias of 3 RCTs was serious [5,10,15],which was classified as high risk.. In the evaluation of outcome data missing and outcome selection reports, one RCT was rated as potential risk [12,13]. There was no blinding method in 12 studies because this type of study could not be blinded to subjects and intervention implementers.

### 3.3 Meta-analysis of results

#### 3.3.1 Body weight index

Ten randomized controlled trials reported weight outcomes, encompassing 464 participants (264 in the experimental group and 200 in the control group). Heterogeneity analysis indicated no significant between-study variability (p = 0.875, I² = 0 %; Fig. 4), and a fixed-effect model was applied. The results of meta-analysis showed that the comprehensive effect of traditional exercise on’ weight’ was WMD = –0.115 (95 % CI: –0.30, 0.07), P<0.01. Subgroup analysis revealed no heterogeneity in the domestic traditional exercise group (p= 0.668, I^2^ = 0 %), and there was no heterogeneity in the foreign traditional exercise group (p= 0.794, I^2^ = 0 %). The heterogeneity test did not reveal significant differences between subgroups (p = 0.608), indicating that the influence of traditional exercises on’ weight’ was similar at home and abroad. Effect estimates indicated no significant effect in the domestic traditional exercise subgroup (z = −1.251, p = 0.211) or in the foreign traditional exercise subgroup (z = −0.343, p = 0.732). The combined effect size (z = –1.192, p = 0.233) showed that traditional exercises had no significant effect on’ weight’. That is, the traditional exercises have no significant improvement in the’ weight’ index of overweight and obese students in the short term (p > 0.05).

**Fig. 4.**
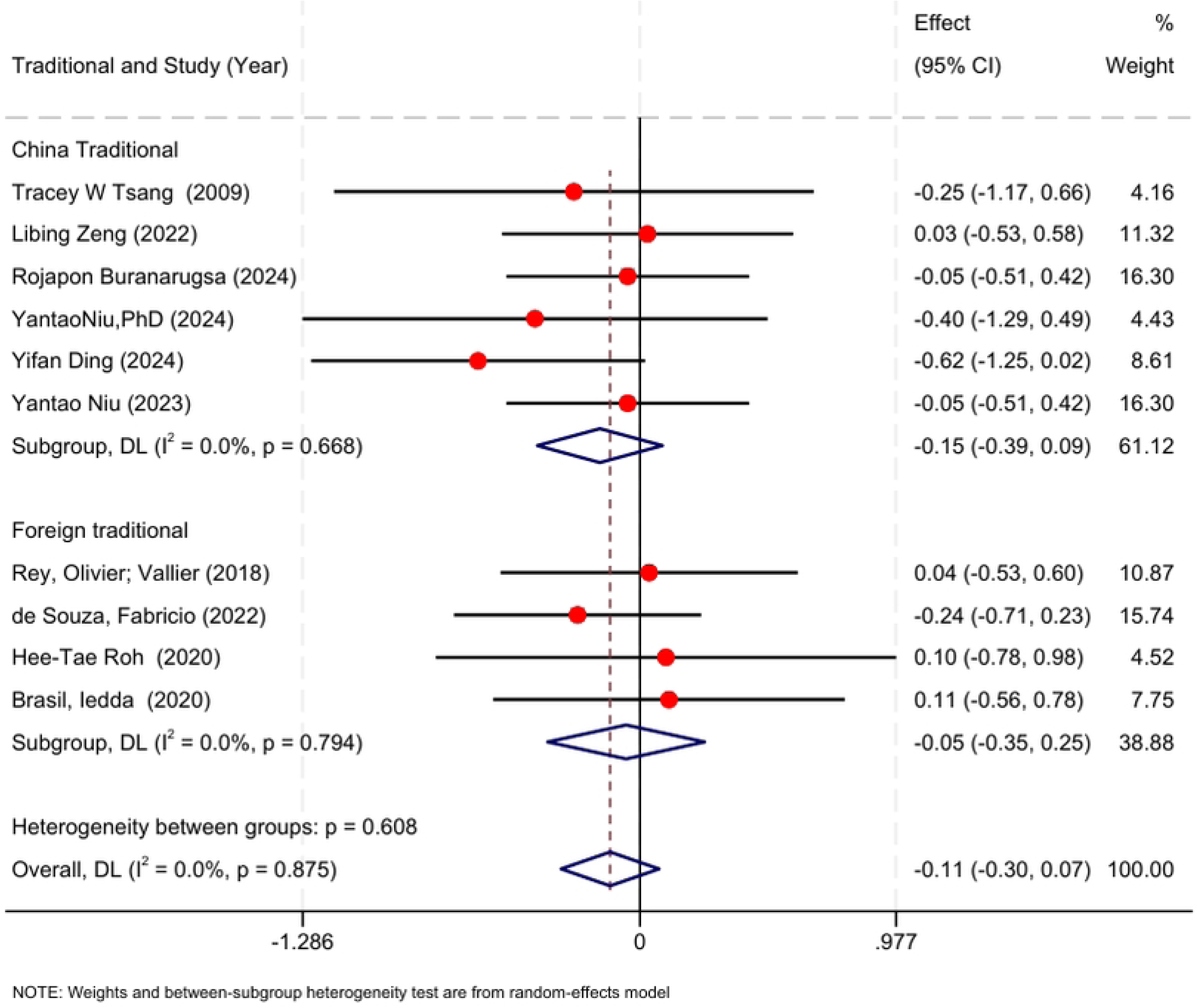
Forest map of the influence of traditional exercises on students’ weight.

#### 3.3.2 BMI indicators

Six randomized controlled trials reported BMI outcomes, encompassing 220 participants (113 in the intervention group and 107 in the control group). Heterogeneity analysis indicated no significant variability among studies (p = 0.576, I² = 0%; Figure 5). The meta-analysis results indicated that the comprehensive effect of traditional exercises on’ BMI’ was WMD = –0.33 (95 % CI: –0.60, –0.06), P<0.01, I^2^=0%. Subgroup analysis showed that there was no heterogeneity in the domestic traditional exercise group (p = 0.441, I^2^ = 0 %), and there was no heterogeneity in the foreign traditional exercise group (p = 0.971, I^2^ = 0 %), and there was no significant difference in the heterogeneity test between the subgroups (p > 0.05). The results of the effect test showed that the domestic traditional exercises significantly improved BMI (z = –2.542, p = 0.011), while the foreign traditional exercises had no significant effect on BMI (z = –0.731, p = 0.465). The combined effect size (z = –2.410, p = 0.016) showed that traditional exercises had a significant effect on BMI, and the improvement effect of Chinese traditional exercises on BMI of overweight and obese students was more significant.

**Fig. 5.**
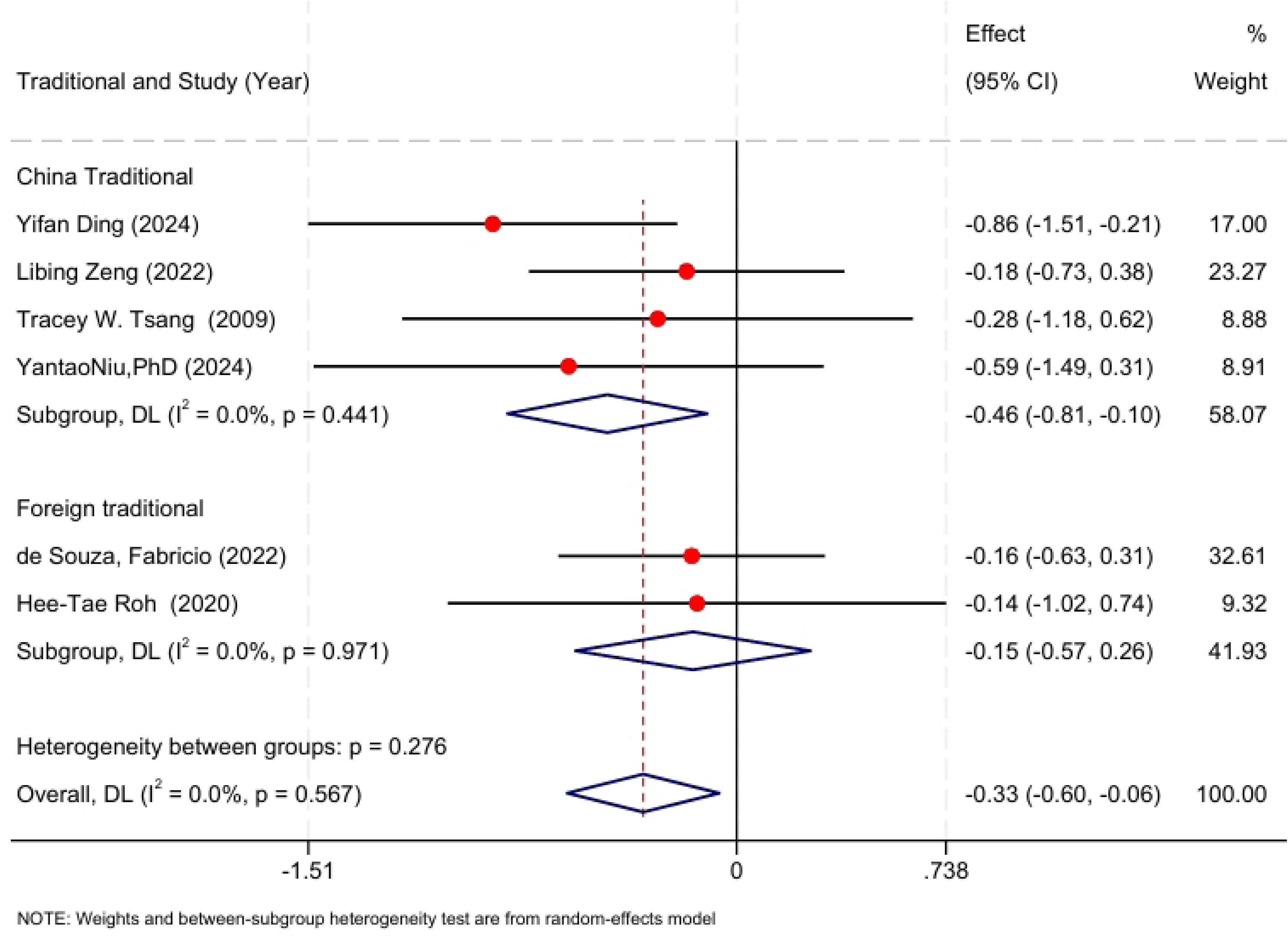
Forest map of the influence of traditional exercises on students’ BMI index.

#### 3.3.3 BF indicator

Seven randomized controlled trials (n=362; experimental=198, control=164) reported body fat outcomes. Heterogeneity was non-significant (p=0.434, I²=0%; Fig. 6), and a fixed-effect model was applied. Meta-analysis demonstrated a pooled weighted mean difference (WMD) of –0.31 (95% CI: –0.52 to –0.10; p<0.01). Subgroup analysis revealed no heterogeneity among domestic traditional exercise studies (p=0.769, I²=0%) and moderate heterogeneity in foreign traditional exercise studies (p=0.152, I²=43.2%). Overall heterogeneity was non-significant (p = 0.434, I² = 0%), and subgroup heterogeneity analysis likewise showed no significant differences (p = 0.782). The effect size test showed that the domestic traditional exercises had a significant effect on reducing’ BF’ (z = –2.087, p = 0.037), while the effect of foreign traditional exercises was not significant (z = –1.524, p = 0.127). The combined effect size (z = –2.911, p = 0.004) showed that traditional exercises had a significant effect on’ BF’, indicating that traditional exercises had a significant improvement effect on’ BF’ indicators of overweight and obese students in the short term (p < 0.05).

**Fig. 6.**
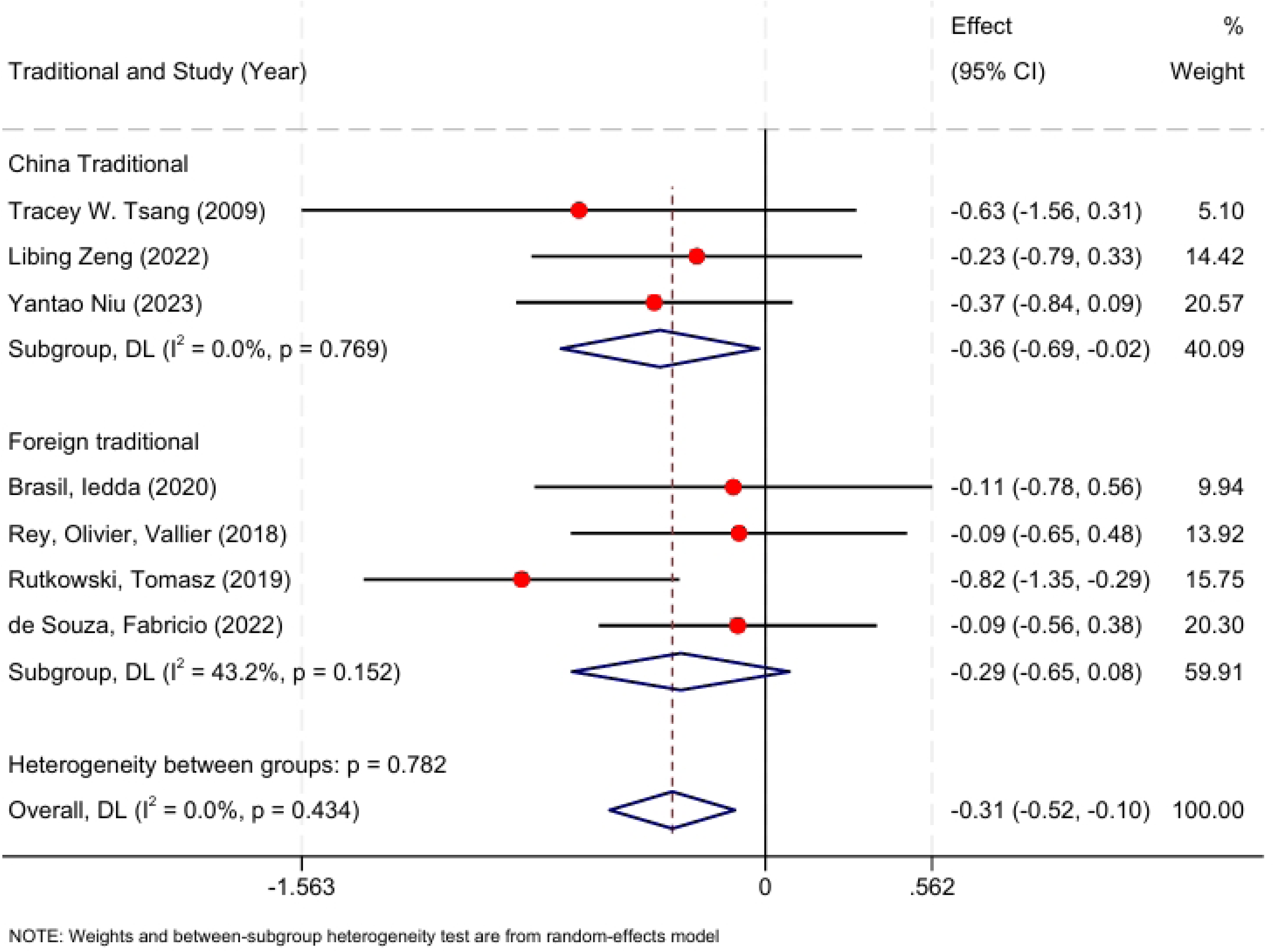
Forest map of the influence of traditional exercises on students’ BF index.

#### 3.3.4 Waist circumference indicators

Three randomized controlled trials assessed waist circumference, encompassing 109 participants (57 in the experimental group and 52 in the control group). Heterogeneity tests were performed on the three included studies (Figure 7), and the results were shown in Figure 7. The pooled results (p = 0.234, I^2^ = 31.2 %) showed low heterogeneity (I²<50%, p>0.05). The results of meta-analysis showed that the comprehensive effect of traditional exercises on waist circumference was [ WMD (95 % CI) = –0.34 ( –0.82,0.13), p < 0.01]. Heterogeneity test showed that there was no significant difference in the effect of traditional exercises on waist circumference [ Chi^2^ = 2.91, df = 2 (p = 0.234), p > 0.05]. Effect size analysis indicated no statistically significant improvement in the’ waist circumference’ index by traditional exercises (z = –1.420, p = 0.156). That is, the traditional exercise method has no significant improvement effect on the’ waist circumference’ index of overweight and obese students in the short term (p > 0.05).

**Fig. 7.**
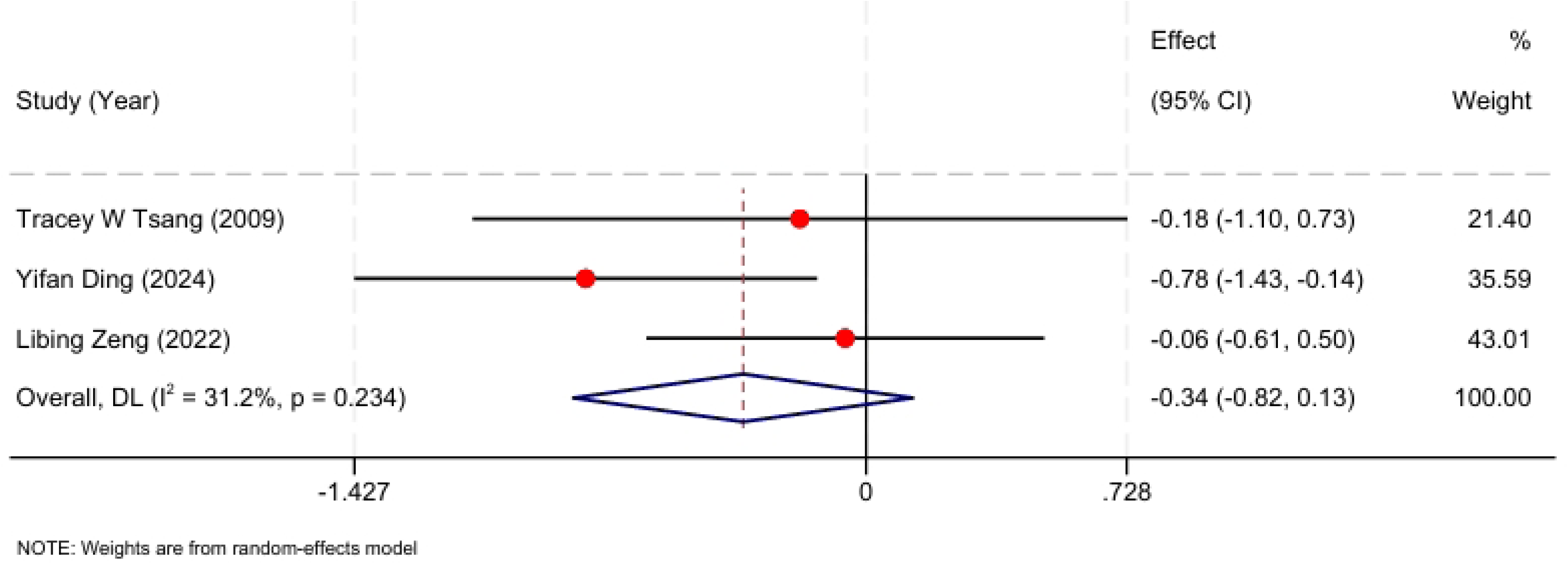
Forest map of the influence of traditional exercises on students’ waist circumference index.

#### 3.3.5 TC indicators

Four randomized controlled trials reported’ TC’ indicators, and 211 subjects were included. The experimental group comprised 122 participants, while the control group comprised 89 participants. Heterogeneity tests were performed on the 4 studies included (Figure 8), and the combined results (p = 0.198, I^2^ = 35.8 %) showed a low heterogeneity (I²<50%, p>0.05). The results of meta-analysis showed that the comprehensive effect of traditional exercises on’ TC’ index was [ WMD (95 % CI) = –0.01 ( –0.37,0.35), p < 0.01]. Heterogeneity analysis indicated no significant differences in the effect of traditional exercises on’ TC’ index [ Chi^2^ = 4.67, df = 3 (p = 0.198), p > 0.05]. The effect size test showed that the traditional exercise method had no statistically significant improvement in the’ TC’ index (z = –0.060, p = 0.952). That is, the traditional exercise method has no significant improvement effect on the’ TC’ index of overweight and obese students in the short term (p > 0.05).

**Fig. 8.**
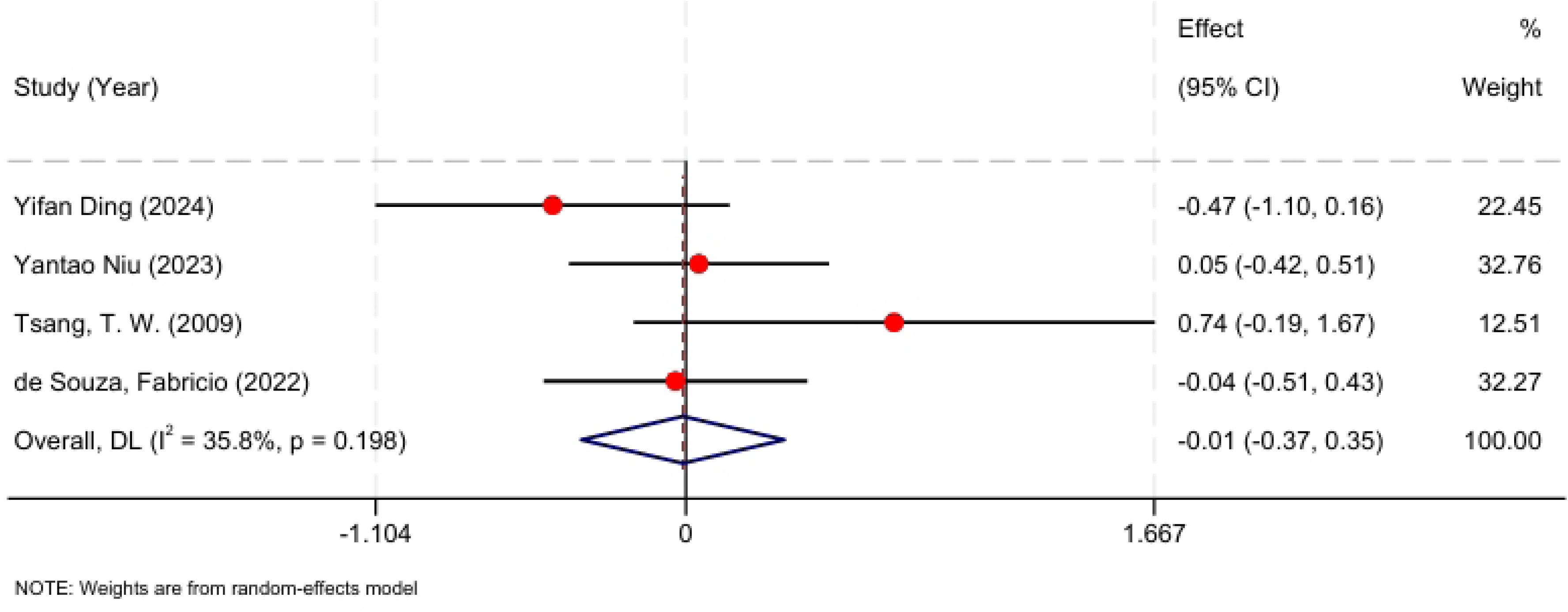
Forest map of the influence of traditional exercises on students’ TC index.

#### 3.3.6 TG Indicators

Three randomized controlled trials assessed triglyceride (TG) outcomes, comprising 129 participants (67 in the experimental group and 62 in the control group). Pooled analysis revealed high heterogeneity (p = 0.005, I² = 80.9%; I² > 75%, p < 0.05; Fig. 9). The results of meta-analysis showed that the comprehensive effect of traditional exercises on’ TG’ index was [ WMD (95 % CI) = 0.33 ( –0.56,1.21), p > 0.01]. Heterogeneity test showed that the effect of traditional exercises on’ TG’ index was significantly different [ Chi^2^ = 10.48, df = 2 (p = 0.005), p < 0.05]. The effect size test showed that the traditional exercise method had no statistically significant improvement in the’ TG’ index (z = 0.726, p = 0.468). That is, the traditional exercise method has no significant improvement effect on the’ TG’ index of overweight and obese students in the short term (p > 0.05).

**Fig. 9.**
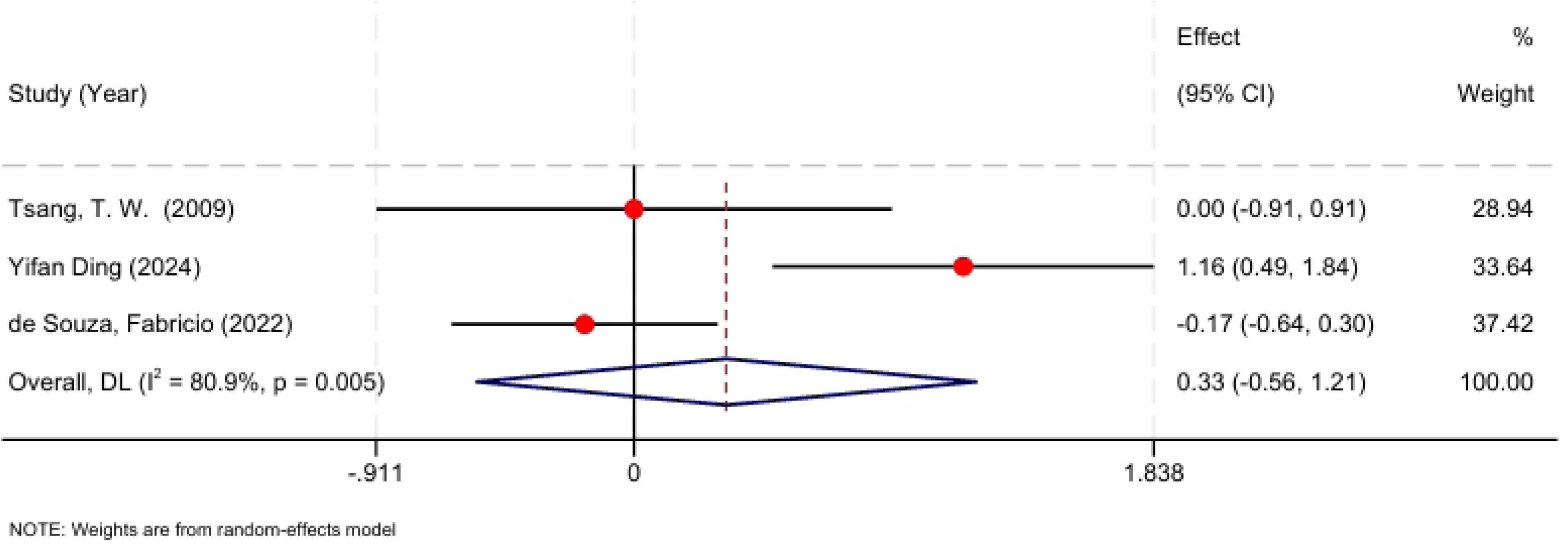
Forest map of the influence of traditional exercises on students’ TG index.

#### 3.3.7 LDL-C index

Three randomized controlled trials reported LDL-C outcomes, encompassing 170 participants (100 in the experimental group and 70 in the control group). Heterogeneity test was performed on the three included studies (Fig.10), and the results were not heterogeneous after merging (p = 0.665, I^2^ = 0 %). The results of meta-analysis showed that the comprehensive effect of traditional exercises on’ LDL-C’ was [ WMD (95 % CI) = 0.03 ( –0.28,0.34), p > 0.01]. Heterogeneity test showed that there was no significant difference in’ LDL-C’ index between traditional exercises [ Chi^2^ = 0.82, df = 2 (p = 0.665), p > 0.05]. The effect size test showed that the traditional exercise method had no statistically significant effect on the’ LDL-C’ index (z = 0.218, p = 0.828). That is, the traditional exercise method has no significant improvement effect on the’ LDL-C’ index of overweight and obese students in the short term (p > 0.05).

**Fig. 10.**
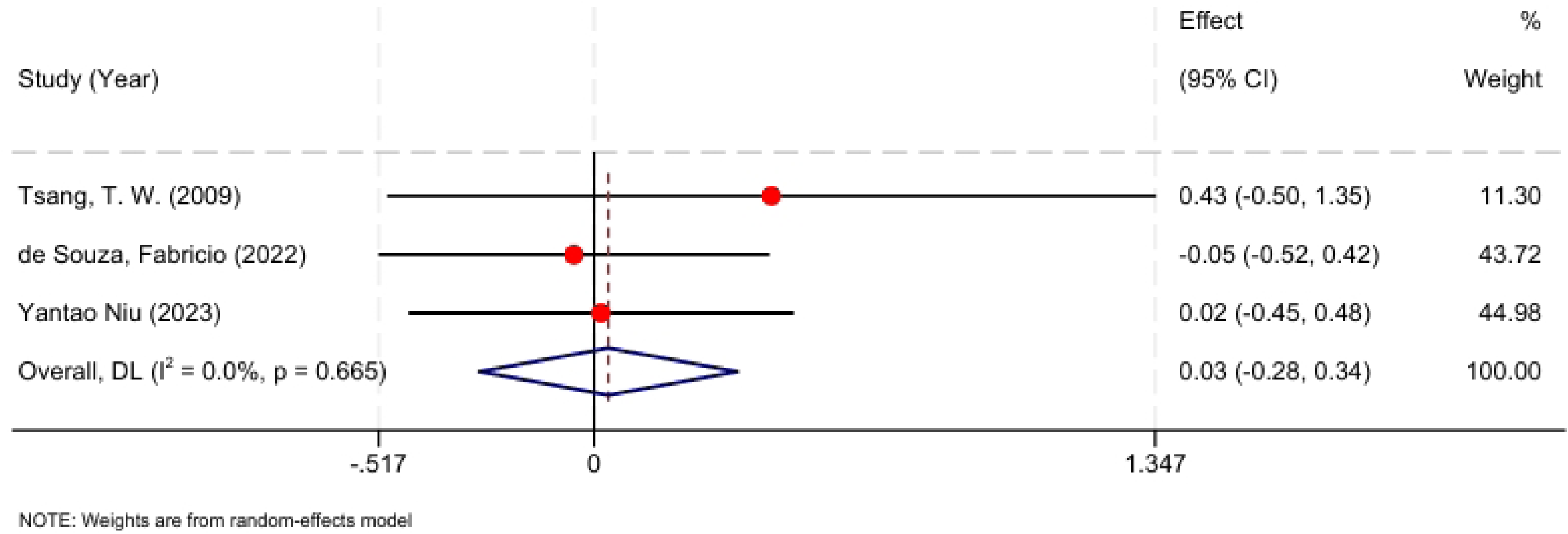
Forest map of the influence of traditional exercises on students’ LDL-C index.

#### 3.3.8 HDL-C indicators

Four randomized controlled trials reported’ HDL-C’ indicators, and 210 subjects were included. The experimental group comprised 121 participants, and the control group comprised 89 participants. Heterogeneity analysis indicated no significant between-study variability (Chi² = 2.57, df = 3, p = 0.464; I² = 0%; Figure 11). Meta-analysis demonstrated a significant effect of traditional exercises on HDL-C, with a pooled WMD of 0.33 mmol/L (95% CI: 0.05 to 0.61; p < 0.01). The effect size test showed that the traditional exercise method had a statistically significant effect on the’ TG’ index (z = 2.319, p = 0.020). That is, the traditional exercise method has a significant improvement effect on the’ HDL-C’ index of overweight and obese students in the short term (p < 0.05).

**Fig. 11.**
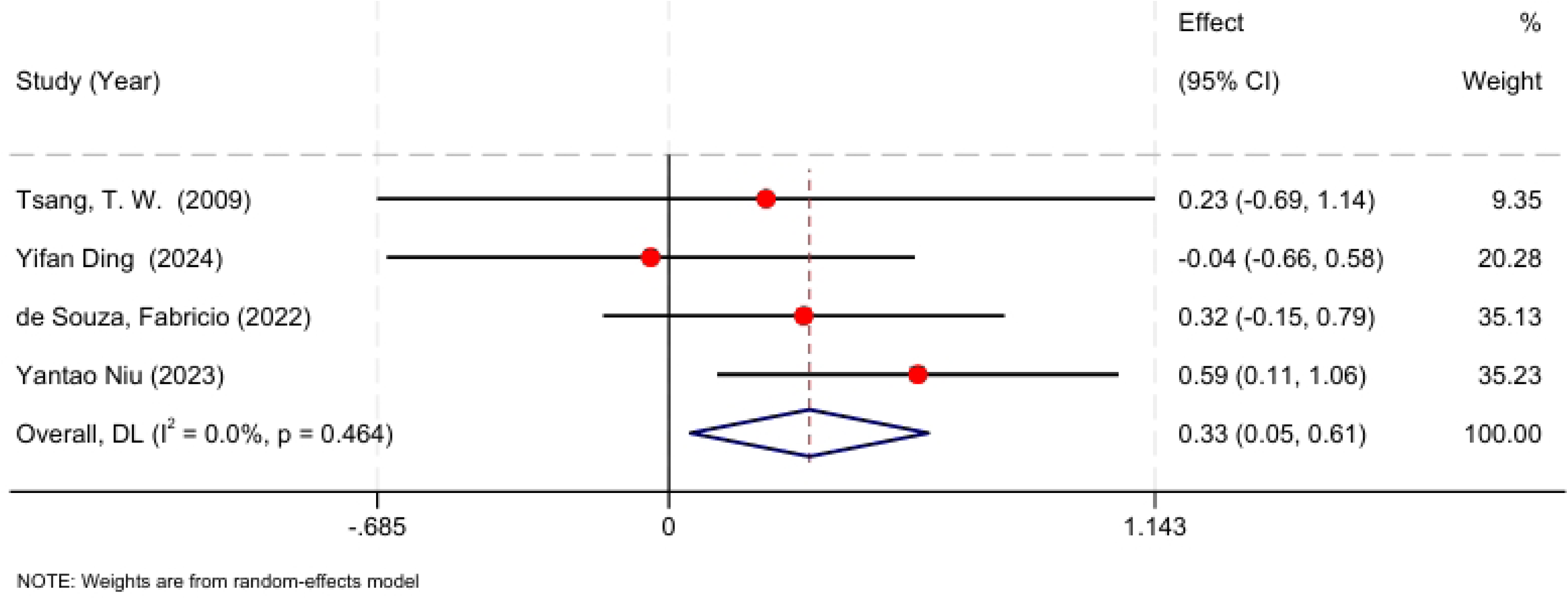
Forest map of the influence of traditional exercises on students’ HDL-C index.

## 4 Discussions

This analysis incorporated twelve randomized controlled trials to assess the impact of traditional ethnic sports on weight reduction and blood lipid profiles in overweight and obese students. A total of 520 overweight or obese students aged 8 to 23 years and BMI > 23.9 were included in the study. The intervention forms included Chinese Wushu, Yang’s Tai Chi, Taekwondo, Boxing, Judo and so on. The results showed that short-term (6-16 weeks) traditional ethnic sports practice significantly reduced BMI (WMD = –0.33, p < 0.05) and body fat rate (WMD = –0.31, p < 0.05), and increased HDL-C (WMD = 0.33, p<0.05). Although the overall weight, waist circumference, TC, TG, LDL-C showed an improvement trend, but did not achieve the statistical significance level. The research shows that national traditional sports have beneficial body composition and blood lipid regulation effects on adolescent overweight and obesity intervention, especially in improving HDL-C.

This study found that the improvement of BMI and body fat was comparable to that of aerobic combined resistance training, and, the observed improvement in HDL-C was consistent with prior meta-analyses, indicating that the duration of exercise intervention rather than type had a more significant effect on HDL-C level [16]. It has been generally believed that significant reductions in LDL-C and TG usually require longer or greater weight loss, while the increase in HDL-C does not depend on weight change. Fan et al.’ s review shows that continuous endurance training can effectively improve blood glucose metabolism, and mixed exercise has more advantages in lipid metabolism optimization [17]. Liu’s study found that exercise patterns had no significant effect on HDL-C, but the duration of intervention was positively correlated with HDL-C improvement [13].. Exercise interventions in overweight and obese students exhibit time-dependent and modality-specific effects on blood lipid profiles. Tsai’s study showed that 12 weeks of Tai Chi training can significantly improve blood lipids, reduce LDL-C and TC, and increase HDL-C [15]. Other studies have also found that at least more than 24 weeks of aerobic combined resistance training can be observed to significantly reduce LDL-C and TG, and increase HDL-C [19–21]. It can be seen that traditional exercise (such as Taijiquan) requires similar intervention duration to achieve the same blood lipid improvement effect as aerobic exercise.

Exercise promotes fat oxidation, glucose transport, improves insulin sensitivity, and increases HDL-C by activating the skeletal muscle AMPK signaling pathway. At the same time, exercise can accelerate triglyceride clearance, up-regulate lipoprotein lipase, reduce serum leptin, increase adiponectin secretion, and inhibit inflammatory factors (such as IL-6, CRP). Martial arts and tai chi practice can also regulate growth hormone and DHEA-S, optimize energy metabolism and lipid balance. The uniqueness of traditional national sports lies in its synergistic effect of integrating’ meaning, qi and shape’, which affects the body’s physiology and psychology. Taking Taijiquan and Qigong as examples, traditional exercises strengthen the regulation of’ qi’ flow and balance internal and external energy through mind training guided by’ meaning’. The slow movement of tai chi combined with mind guidance helps to dredge meridians, reconcile qi and blood, improve microcirculation and tissue metabolism [22]. In addition, the’ shape’ exercises in national traditional sports, such as standing, horse walking, leg raising, etc., combined with isometric and dynamic stretching movements, exercise core and deep muscles, improve muscle strength and flexibility, while low impact, systemic participation, can effectively improve metabolic needs [23–25]. Compound movements also promote tendon and fascia remodeling, improve energy storage and release, and promote fat oxidation and muscle formation [25–27]. In addition, Tai Chi’s abdominal deep breathing and movement coordination mobilize the parasympathetic nerve, reduce sympathetic nerve excitement, and reduce cortisol secretion. Evidence indicates that Tai Chi practice enhances heart rate variability, lower blood pressure, reduce psychological stress and anxiety, thus contributing to weight control and blood lipid improvement [28,29].

Secondly, martial arts routines, Taiji’s artistic conception and Duanwei promotion system can enhance learning interest and motivation, and ensure the intensity and duration of intervention. Psychologically, the practice of’ unity of form and spirit’ improves self-efficacy, helps adolescents gain a sense of achievement in the process of mastering movements, and enhances persistence. In the social and cultural mechanism, the cultural identity and Duan system of Wushu routines, the collective performance form of Tai Chi, and the long-term compliance through group identity and goal motivation. Compared with high-intensity training (HIIT), traditional ethnic sports show better continuous participation rate in adolescent obesity intervention due to low fatigue and high interest. The research shows that the social identity in sports enhances the collective motivation and continuous participation motivation. National sports can form a common identity in the crowd through cultural inheritance and reduce the risk of abandoning training in the middle [30]. Combat traditional sports (such as kung fu, taekwondo, karate) combined with high-intensity training such as fighting and wrestling, can improve cardiopulmonary endurance and muscle strength in a short time. Studies have shown that adolescent kung fu training can effectively promote energy metabolism, fat oxidation and body composition improvement [31]. These combined effects of physical and mental integration make traditional ethnic sports have unique physiological and psychological advantages in the intervention of overweight and obesity.

Subgroup analysis found that domestic traditional exercises (such as Tai Chi and Ba Duan Jin) had a significant effect on improving BMI and body fat rate: BMI, WMD = − 0.48 (p = 0.004), body fat rate WMD = − 0.42 (p = 0.003); although the foreign subgroup showed a negative trend, it was not significant (p > 0.05). This shows that local exercises are more effective in improving body composition. Nanquan (NQ) training in martial arts reaches 83.1 % of the maximum heart rate at 25-40 seconds, and the energy consumption per unit time is higher than that of conventional aerobic exercise [32]. Tai Chi, a low-intensity aerobic exercise, emphasizes respiratory cooperation and indirectly promotes fat metabolism by improving cardiopulmonary function. When Tai Chi is combined with nutritional intervention, the improvement of body composition is more obvious [33]. The latest research shows that Baduanjin has better fat-reducing effect, and Tai Chi is more effective in improving waist-to-hip ratio, which is the best way to reduce fat while increasing muscle [34]. Localized exercises have a more significant effect on the improvement of body composition due to higher exercise intensity, multi-muscle complex exercise design, respiratory idea coordination and cultural adaptability. In the overall analysis, most of the outcome indicators (body weight, BMI, body fat rate, HDL-C) had low heterogeneity, but triglyceride (TG) showed high heterogeneity (I^2^ = 80.9 %, p = 0.005). The sensitivity analysis showed that the change of TG was affected by many factors such as the duration, frequency and region of intervention, which may be related to the small sample size, the difference of intervention plan and the difference of dietary control.

In this study, RCT data from China, Australia, Europe and South America were collected, and WMD indicators were used to find that the effects of different sports and populations were consistent, which enhanced the robustness of the conclusions. The research covers high-intensity combat projects (such as kung fu, taekwondo) and low-intensity physical and mental adjustment projects (such as Yang’s Tai Chi, Bafa Wubu Tai Chi), which comprehensively reflects the impact of different exercise modes on overweight and obese students. The study examined alterations in body composition and blood lipid profiles in relation to underlying physiological mechanisms such as AMPK activation and fat oxidation, as well as psychological mechanisms such as cultural belonging and self-efficacy, and clarified the advantages of traditional ethnic sports. Subgroup analysis showed that local exercises had more advantages in improving BMI and body fat rate, providing empirical evidence for obesity intervention programs. The limitations of this study include large differences in intervention types and doses, and small sample size, especially in the analysis of blood lipid indicators such as TG and LDL-C, with significant heterogeneity. In addition, data on long-term weight maintenance remain insufficient and blood lipid improvement. Future studies should standardize exercise dose, verify long-term effects and analyze the mechanism in depth. Through multi-center large sample studies, the dose effects of different traditional sports should be compared, and longitudinal follow-up studies are needed to assess the durability of weight maintenance and the sustained improvements in blood lipid profiles.

## 5 Conclusion

This study confirmed the effectiveness of traditional ethnic sports in overweight and obese students through Meta-analysis of 12 RCT studies. National traditional sports significantly reduced BMI and body fat rate, and increased high-density lipoprotein cholesterol level. However, no significant short-term changes were observed in body weight, waist circumference, total cholesterol, triglycerides, or low-density lipoprotein cholesterol. Subgroup analysis revealed that indigenous Chinese exercises produced more pronounced reductions in BMI and body fat percentage. The overall heterogeneity of the research results is low and the results are robust, but the heterogeneity of TG is high, suggesting that future research needs to unify the intervention dose and dietary control, expand the sample size, and further explore the role of the full spectrum of blood lipids. In general, national traditional sports can be used as an exercise intervention program for overweight and obese students, and has good promotion value. In the future, long-term follow-up studies and multi-center large sample verification should be combined to further optimize the intervention plan and clarify the dose-effect relationship.

## Data Availability

All relevant data are within the manuscript and its Supporting Information files.

## Notes

### Competing Interest Statement

The authors have declared that no competing interests exist.

### Funding Statement

The author(s) received no specific funding for this work.

